# A topic analysis of traditional and social media news coverage of the early COVID-19 pandemic and implications for public health communication

**DOI:** 10.1101/2020.07.05.20146894

**Authors:** Wallace Chipidza, Elmira Akbaripourdibazar, Tendai Gwanzura, Nicole M. Gatto

## Abstract

Knowledge gaps may initially exist among scientists, medical and public health professionals during pandemics, which are fertile grounds for misinformation in news media. We characterized and compared COVID-19 coverage in newspapers, television, and social media, and discussed implications for public health communication strategies that are relevant to an initial pandemic response. We conducted a Latent Dirichlet Allocation (LDA), an unsupervised topic modelling technique, analysis of 3,271 newspaper articles, 40 cable news shows transcripts, 96,000 Twitter posts, and 1,000 Reddit posts during March 4 - 12, 2020, a period chronologically early in the timeframe of the COVID-19 pandemic. Coverage of COVID-19 clustered on topics such as epidemic, politics, and the economy, and these varied across media sources. Topics dominating news were not predominantly health-related, suggesting a limited presence of public health in news coverage in traditional and social media. Examples of misinformation were identified particularly in social media. Public health entities should utilize communication specialists to create engaging informational content to be shared on social media sites. Public health officials should be attuned to their target audience to anticipate and prevent spread of common myths likely to exist within a population. This will help control misinformation in early stages of pandemics.

## Introduction

On December 31, 2019, the World Health Organization (WHO) was alerted to the occurrence of a series of cases of pneumonia with unknown etiology in Wuhan City, China, which were subsequently linked to exposures at a seafood and live animal market (World Health Organization, 2020). Chinese researchers identified the cause of the disease to be a new type of coronavirus (Ren et al., 2020), later named COVID-19 by the WHO(World Health Organization, 2020). Between January and mid-March, the disease spread from its epicenter to other Chinese cities and to over 150 countries across all continents (Johns Hopkins University & Medicine, 2020). On March 11, the WHO declared COVID-19 a pandemic(World Health Organization, 2020), and by April 29, the number of confirmed cases globally exceeded 3 million, with 1/3 of these identified in the United States (US) (Johns Hopkins University & Medicine, 2020).The pandemic wrought havoc on public health and medical systems internationally, caused severe disease and death among those infected overwhelming medical clinics and hospitals; resulted in closures of schools, and cancellation or postponement of sporting and entertainment events; led to restrictions in travel and overall disruptions of daily life, and upended global financial markets(2020).

In emergency situations, communication of important information to affected populations is critical (US Department of Health and Human Services Centers for Disease Control and Prevention, 2018 Edition; World Health Organization, 2017). Information from governments, public health and medical entities during pandemics is vital to decision-making (Pan American Health Organization, 2009), to taking actions to contain existing disease, and to prevent its further spread. A reliance on the news media for communication is an expected and deliberate component of a response to pandemics (US Department of Health and Human Services Centers for Disease Control and Prevention, 2018 Edition). In the US, people seek and receive news information from numerous different sources, including newspapers, radio, television, and increasingly, social media (Shearer, 2018, December 10). During outbreaks of novel infectious diseases, an understanding of the disease builds with time and thus initial knowledge gaps may exist among scientists, medical and public health professionals. These gaps are fertile grounds for spread of misinformation in news media sources (Lee & Basnyat, 2013). For traditional media, journalists may misreport information, from misunderstanding scientific facts, receiving wrong information, or through sensationalized reporting (Larsson et al., 2003). For social media, users can participate semi- or fully anonymously and spread false information without fear of repercussions (Kling et al., 1999). The prevalence of misinformation could itself be a source of risk in pandemic situations (Smallman, 2015), with numerous options for news sources presenting a challenge for communication. Further, topics emphasized or underemphasized by media potentially pose risks to the public; for example, the media may lend credibility to unproven cures, or underreport ways to prevent spread of the disease. A question arises as to whether more effective strategies with news media would help to achieve public health objectives towards prevention and control in pandemic situations like COVID-19.

A variety of studies have previously examined media coverage of pandemics. A content analysis of British media coverage of SARS in 2003, concluded that media tended to characterize SARS as of Chinese origin, and conveyed that the superiority of “Western scientific biomedicine” would contain its spread (Washer, 2004). An investigation of how the H1N1 pandemic was portrayed found corporate organizations including the National Pork Producers Council and National Pork Board adopted a more reassuring tone in response to the crisis than governmental organizations such as the Centers for Disease Control and Prevention (CDC) and the Department of Health and Human Services (Liu & Kim, 2011). Other researchers faulted the WHO and CDC’s response to the H1N1 pandemic for enabling stigmatization of “Mexicans and other Latinos in the US … as carriers of the virus”(McCauley et al., 2013). A mixed methods study of Dutch media coverage of H1N1 implicated both media *and* expert sources for overstating the threat of the virus(Vasterman & Ruigrok, 2013). The majority of existing research on media coverage of pandemics to date have relied on qualitative methods for analysis. As the volume of information accumulating on the internet on news topics expands, more sophisticated methods of analyses are needed.

The value of machine learning techniques such as Latent Dirichlet Allocation (LDA) in understanding health-related discussions on social media has been previously demonstrated. Data posted on social media has been used to aid disease surveillance during the 2011 *Escherichia coli* outbreak in Germany (Diaz-Aviles et al., 2012), the 2010 influenza epidemic in the US (Achrekar et al., 2011), and the 2009 H1N1 pandemic (Al-garadi et al., 2016). Topic analysis has been employed to discover major health and disease topics of interest discussed on Twitter(Paul & Dredze, 2014). Further, in online healthcare communities, users generally post about symptoms, exams, medications, procedures, and complications relating to critical illnesses (Lu et al., 2013). While previous research shows the growing importance of social media in responding to emerging health crises, studies are needed on the nature of pandemic-related information. The flow of information on social media originates initially within smaller, specific subcommunities before spreading to a wider audience on the internet (Weninger et al., 2013). Coverage of pandemics is not restricted to health-related information like symptoms and treatments(Vasterman & Ruigrok, 2013). Furthermore, social media has become pervasive in everyday life, giving a voice to people on important issues.

COVID-19 may be the most disruptive international health issue in modern times, and is dominating news media. This paper explores the nature of COVID-19 coverage in traditional news and social media. Our objectives are first, to characterize the nature of information received by news consumers of newspapers, television, and social media using Latent Dirichlet Allocation (LDA) without making any *a priori* hypotheses, in order to discover topics associated with COVID-19 on different platforms. Our second objective is to compare topic configurations across platforms to examine potential differences. Finally, based on our observations, we discuss implications for communication strategies by public health entities that are relevant to an initial pandemic response.

## Materials and Methods

### Topic Modeling Using Latent Dirichlet Allocation

LDA, an unsupervised machine learning technique (Blei et al., 2003), is an exploratory algorithm useful for discovering underlying topics within large bodies of text commonly referred to as a corpus. LDA is a generative probabilistic model in that it simulates the random process by which a given document within the corpus could have been generated (Blei, 2012). This inductive approach identifies topics that one might not anticipate. LDA has been shown to perform better than other topic modeling techniques in health-related text mining (Sarioglu et al., 2012). The goal of LDA is to compute the posterior probability given evidence, i.e. the conditional distribution of topics given documents within the corpus (Blei, 2012). Calculating this posterior probability requires computation of the joint probability distribution of β, θ, and *z* across all *w* (Figure 1) and dividing it by the probability of observing the corpus across all possible topic models. Algorithmically, the procedure begins with random guesses of β and θ and a pre-specified number of topics *K*. Each word in a document is randomly assigned to a topic, and this process repeats conditioned on the current topic distribution. A word is reassigned to another topic if the topic rarely appears within the document, or if the word rarely appears in the current topic. The algorithm converges when there are no new reassignments, or when the number of iterations is reached, resulting in a per document topic distribution and per topic word distribution for a corpus.

**Figure 1:**
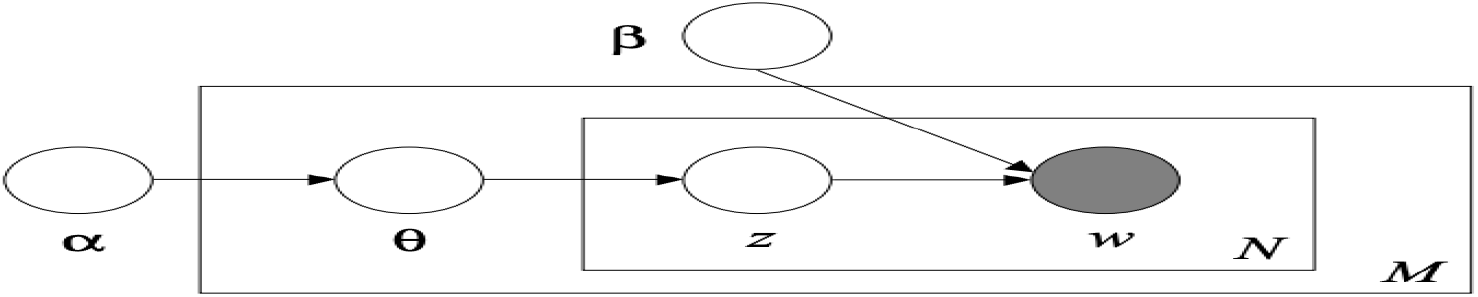
LDA in plate notation (adapted from Blei et al. 2003) with parameters: *α*-initialization parameter controlling the per document topic distribution. β - per topic word distribution. θ - per document topic distribution *N* - the inner plate denoting the words contained in a given document *M* - the outer plate denoting the documents constituting the corpus *w* - specific word in a given document. It is the only observed variable in the model. *z* - the topic assignment for a specific word within a document.

### Data Collection and Analysis

Data from newspaper, television, and social media sources (Table 1) were obtained for the nine-day period of March 4 - 12, 2020, selected for being early in the pandemic. We employed searches with keywords “coronavirus” and “COVID” in bodies of Twitter posts, newspaper articles, and television transcripts. The same keyword searches were used for titles in Reddit submissions, specifically the r/all subreddit which aggregates the most popular submissions across the Reddit community. Raw data were collected and saved in text files for analyses (Figure 2).

**Table 1:**
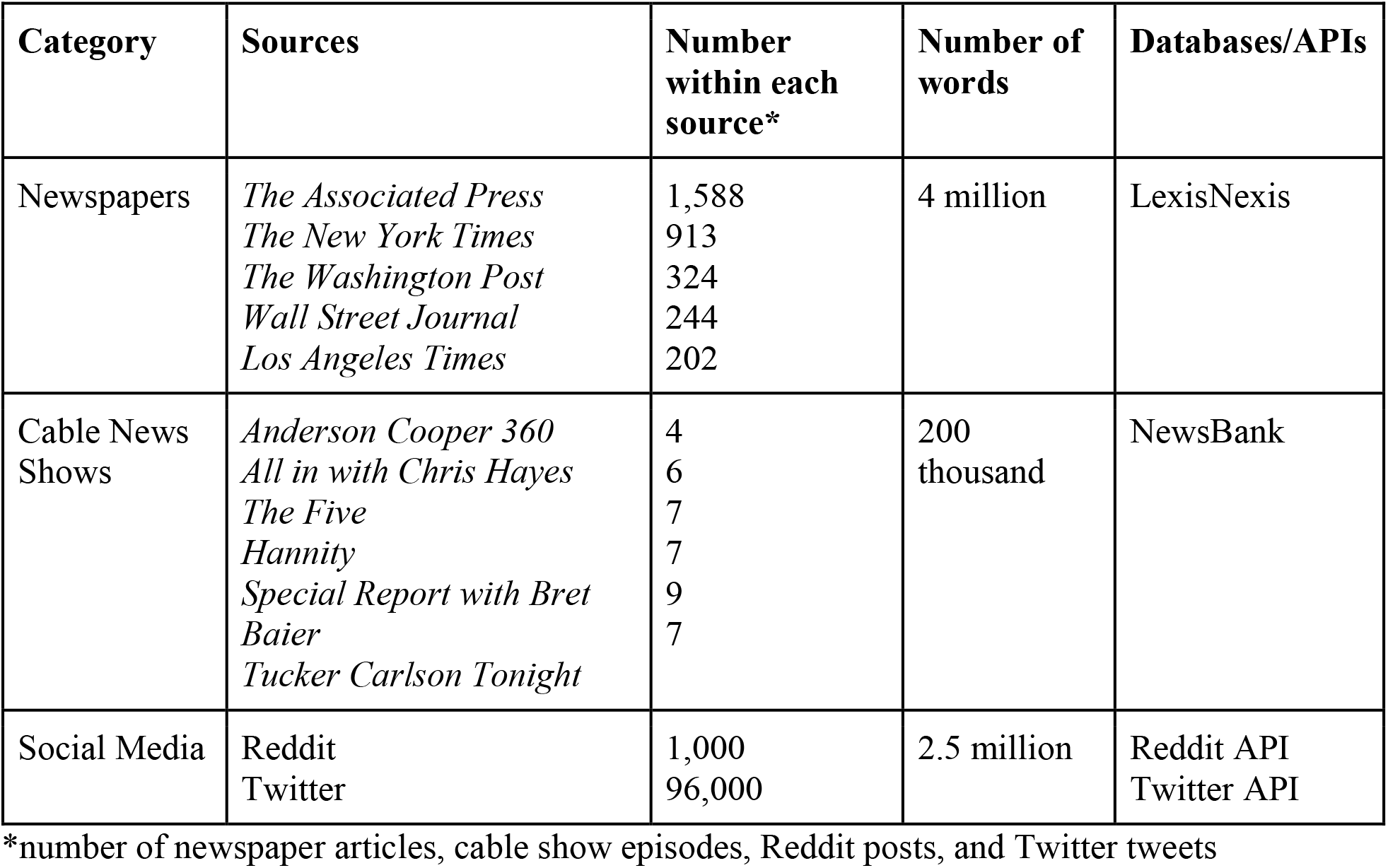
Data sources and number of each source, words and databases employed in news media search for LDA analysis.

| Category | Sources | Number within each source* | Number of words | Databases/APIs |
| --- | --- | --- | --- | --- |
| Newspapers | *The Associated Press*<br>*The New York Times*<br>*The Washington Post*<br>*Wall Street Journal*<br>*Los Angeles Times* | 1,588<br>913<br>324<br>244<br>202 | 4 million | LexisNexis |
| Cable News Shows | *Anderson Cooper 360*<br>*All in with Chris Hayes*<br>*The Five*<br>*Hannity*<br>*Special Report with Bret Baier*<br>*Tucker Carlson Tonight* | 4<br>6<br>7<br>7<br>9<br>7 | 200 thousand | NewsBank |
| Social Media | Reddit<br>Twitter | 1,000<br>96,000 | 2.5 million | Reddit API<br>Twitter API |

\*number of newspaper articles, cable show episodes, Reddit posts, and Twitter tweets

**Figure 2:**
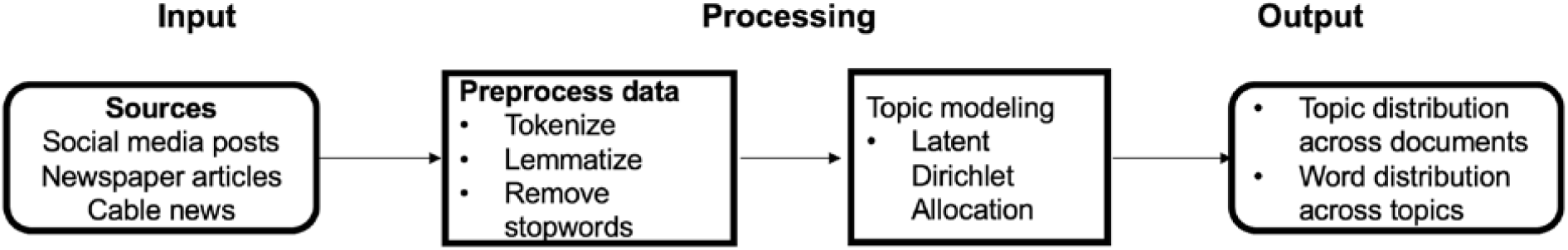
Data collection and analyses process, LDA analysis of news media.

Topic analysis requires preprocessing of raw data to a specific format (Zhao et al., 2011). We used the process of lemmatization to convert individual words to their root word. Stopwords which occur very frequently in the English language bring noise into the topic discovery process, so were similarly removed. We employed term frequency*-*inverse document frequency (TF-IDF) to derive the most important words for each corpus(Ramos, 2003). TF-IDF is a term weighting scheme that allocates higher importance for a word appearing frequently within a document, while controlling for the word’s appearance across all documents.

### Understanding topic modeling output

Consistent with the spirit of LDA(Blei, 2012), we prioritized human interpretability of resultant topics and settled on *K*=3, to ensure adequate coverage of interpretable topics across traditional and social media (DiMaggio et al., 2013). Having retrieved the top words defining each of the topics across corpora, we scored individual components of each corpus against generated topics, and ranked them to identify which topics defined each component. Two authors (WC & NMG) derived labels based on content loading highest on individual topics, and then discussed to resolve any differences. This methodological approach is consistent with previous research employing topic modeling(DiMaggio et al., 2013). IRB approval was not obtained as the research did not involve human subjects.

## Results

The top words defining coverage of COVID-19 across traditional and social media over the selected time period were *case, trump, travel, virus, china, world, test, health*, and *public*. Topics discovered varied based on the category of the media source (Table 2) and individual articles were frequently reflected in more than one topic. For example, an article about a football team cancelling an autograph session citing fears of the virus loaded on the global topic (which included the *sports* term) in the newspaper corpus, but was also a mixture of the epidemic and economy topics.

**Table 2.** Discovered topics across traditional and social media corpora with top words in topic and top words across corpora.

| Source | Topic label | Top words in topic* |
| --- | --- | --- |
| Newspapers | **Epidemic** | <u>health</u>, <u>virus</u>, disease, <u>trump</u>, state, type, outbreak, <u>travel</u>, <u>case</u>, business, administration, home, italy |
|  | **Economy** | <u>public</u>, <u>china</u>, industry, company, economic, market, care, infectious, presidential, word |
|  | **Global** | government, sports, europe, <u>world</u>, <u>test</u>, stock, load |
| Cable News | **Testing** | <u>test</u>, travel, important, medical, fact, economic, pence, risk |
|  | **Politics** | <u>trump</u>, <u>health</u>, <u>china</u>, <u>case</u>, state, <u>public</u>, american, <u>world</u>, white, doctor, fauci, government |
|  | **Consequences** | <u>virus</u>, number, home, work, administration, disease, market, states, crisis |
| Twitter | **Politics** | <u>trump</u>, breaking, positive, hanks, <u>public</u>, <u>testing</u>, <u>health</u>, monkey, response, <u>virus</u>, travel |
|  | **Pandemic** | free, gather, italy, house, covid19, pandemic, outbreak, basketball, director, porter, barackobama, holy |
|  | **Testing/Response** | <u>test</u>, <u>case</u>, <u>world</u>, <u>china</u>, spread, source, overreaction |
| Reddit | **Epidemic** | <u>virus</u>, <u>test</u>, covid, meet, <u>china</u>, situation, <u>trump</u>, risk, <u>world</u>, government, travel |
|  | **Society** | home, <u>case</u>, work, school, f**k, <u>public</u>, care, part |
|  | **Consequences** | sick, <u>health</u>, number, s**t, state, life, spread, company, reason, italy, plan |

\*words appearing across all corpora indicated using underlined font

### Newspapers

The “epidemic” topic was comprised of terms relating to the *disease outbreak* and its spread. An article in the *Wall Street Journal* which loaded on this topic reported that at least 100,000 people worldwide were infected with the virus, and estimated a case fatality rate of 2-4%. An *Associated Press* article explained how to distinguish between flu and COVID-19 symptoms. Citing a medical doctor and the WHO, it explained that while flu symptoms (e.g. high fever, extreme exhaustion) manifest suddenly, symptoms for COVID-19 appear more gradually and include dry coughs and shortness of breath. Another article loading highly on this topic from the *New York Times* advocated parents not inform their 6-year-old children about the virus to prevent unnecessary anxiety.

The “economy” topic illustrated economic effects of the pandemic including reduced revenues for various companies due to lower economic activities, bankruptcy of airlines, and efforts of US legislators to increase funding. The third topic demonstrated the global impact of the pandemic with content in articles underscoring effects of COVID-19 beyond the US. As examples, the South Korean government opposition to Japan’s decision to quarantine South Korean visitors; a France-Ireland rugby match was postponed to prevent coronavirus from spreading; and a European and Russian joint mission to launch a rover to Mars was postponed because of COVID-19-related travel restrictions.

### Cable News

The “testing” topic included coverage of the cruise ship off the California coast, and the Vice President’s announcement that passengers would be tested and quarantined if necessary. On multiple programs, cable news personalities advocated the need for more coronavirus testing in the US, and South Korea was noted as an example of how to aggressively test to understand the scope of the outbreak. One commentator noted the country’s overall mitigation strategies, including the number of tests conducted each day and different treatments being tried including Plaquenil and chloroquine. (*Tucker Carlson Tonight*, 3/12/20). Expedited approval of testing kits by the Food and Drug Administration under the emergency use provision was highlighted by some cable news hosts (*Anderson Cooper 360*, 3/5/20).

The “politics” topic included coverage highlighting four members of the US Congress who had self-quarantined after possible contact with COVID-19 positive individuals (*All In With Chris Hayes*, 3/9/20). Some hosts critiqued China’s actions in responding to the outbreak, with one claiming that it might “shut off the supply of pharmaceuticals” to the US (*Tucker Carlson Tonight*, 3/11/20). The same host also criticized the WHO recommendation to refer to the virus by its scientific name, insisting that it should be referred to by its place of origin. The third topic underscored the severe effects of the pandemic. When commentators talked about the *virus*, they also tended to talk about *home* and *work* life. Some linked the virus to disruptions of the supply chain, school closures, increased unemployment, and increased need of telemedicine and working from home. Discussions centered around the prospect of the healthcare system being overwhelmed by the number of patients suffering from COVID-19 disease. Discourse also brought attention to the global economic devastation wrought by the pandemic. The possibility that the impact on the US economy would have implications on the 2020 presidential election was also highlighted.

### Twitter

Based on terms contained in the first topic, associated tweets emphasized how politics and public policy should address the pandemic. Representative tweets which loaded highly on this topic included “Once a vaccine for coronavirus is developed it should be free.” The second topic underscored the global nature of the crisis. Representative tweets included from the WHO: “Of the 118000 #COVID19 cases reported globally in 114 countries more than 90 percent of cases are in just four countries…” The third topic emphasized the need for testing and the backlash to actions required to slow spread of the virus. Tweets included “Severe shortage of tests blunts coronavirus response Boston doctors say,” and “The global pandemic of our time isn’t the Coronavirus. The global pandemic of our time is FEAR”. This topic demonstrates concerns regarding lack of testing capacity in the US, and shows that a significant number of people worried that authorities were overreacting while themselves not taking the pandemic seriously.

### Reddit

Based on terms defining the first topic, posts reflected information-seeking behavior on the part of users wanting to know more about COVID-19. Comments were in response to posts with titles such as “My father is 76 and still works as a doctor… How can my mom and me convince him to stop working?” and “Is corona virus like chicken pox…you get it once and done or can you get it over and over again?” There was substantial discussion on whether fatalities varied by age; the consensus was that for young children and healthy adults, the disease was not “that dangerous.”

The second topic emphasized the impact of COVID-19 on everyday life, with references to *home, work*, and *school* life. Comments loading highly on this topic alerted of the possibility of school closures, working from home, and also staying indoors for an indefinite period. Other users referenced restaurants limiting their hours of service, staying home with children, and hoarding of supplies like food and toilet paper. The third topic articulated serious consequences of COVID-19 with references to severity of illness among patients, sports leagues in various countries canceling their seasons, and people losing jobs and incomes. Some users referenced plans for freezing mortgage and rent payments. Others were not convinced that the US government’s response to the pandemic was adequate.

## Discussion

Our analysis of media sources during March 4-12, 2020 showed that major discovered topics were not predominantly health-related, which would indicate less of a presence of public health entities in news coverage and social media. While there was substantial mention of the presidential administration, references to ventilators, social distancing, and hygiene were more limited. This lack of a prominent public health “voice” represents a missed opportunity for public health and medical experts to establish themselves as trustworthy and credible sources for information and instructions. The nine-day period selected for analysis was relatively early in the US coronavirus pandemic timeline. During this period there were 149 confirmed cases which increased to 1,663, and eleven deaths which increased to 40 (Johns Hopkins University & Medicine, 2020). (For comparison, ten days later on March 22 there were 33,276 confirmed cases and 417 deaths.) Significant news events during the nine-day period included the CDC’s removal of previous restrictions on coronavirus testing, and the declaration of COVID-19 a pandemic by WHO(CNN Editorial Research, 2020). The latter also marked the US announcement of new travel restrictions from Europe. In retrospect, the need for an authoritative voice of health to emerge during this early period was still very much unfilled. An early deficient public health presence creates opportunities for other less informed voices to dominate, or worse, to disseminate misinformation(Mian & Khan, 2020; Pan American Health Organization, 2009; World Health Organization, 2017). This was seen in cable news content where hosts downplayed the risk of infection. A minimized presence of public health in the media could consequently be detrimental to the public being provided with educational information important to reduce spread of the virus. A limitation of our approach is that we selected a specific time frame to conduct our analyses, which was relatively early in the pandemic. Continued analysis of media coverage of the pandemic could suggest additional interpretations. Further, to understand coverage on social media, we analyzed Twitter and Reddit content; future research could explore coverage on other platforms such as Facebook and Instagram.

Pandemic preparedness should include communication plans which are ready and can be activated during early days of the disease’s entry and spread into a population(Pan American Health Organization, 2009; World Health Organization, 2017). Part of preparedness could include some ready-made general informational and educational materials that could be quickly deployed, or at a minimum, templates should be available to facilitate their rapid development and publishing. For COVID-19, early indications of a novel coronavirus causing respiratory illness could have prompted public health entities to release such readied educational materials on covering coughs and sneezes. Our analysis showed instances of myths being perpetuated in social media (e.g., smoking making lungs inhospitable to coronavirus). Materials that are poised for use should anticipate common myths likely to exist within a population. This should be possible if public health entities are well-acquainted with their target audience in advance of a pandemic, as is recommended(Pan American Health Organization, 2009; World Health Organization, 2017).

The perceived unknown and uncontrollable nature of the novel coronavirus and potential for severe disease resulting from its infection would place the pandemic in the low familiarity/high dread of Slovic’s psychometric risk paradigm (2000). This serves to indicate some of the challenges (Vaughan & Tinker, 2009) underlying risk communication. The public’s perception of risk may be shaped early in a pandemic and may be difficult to change once formed(Sheppard et al., 2012). Together these emphasize the importance for public health entities to seize early communication opportunities and follow a well-conceived communication plan. Initial communication from public health during a novel infectious disease outbreak should acknowledge the uncertainty involved with the newly identified agent and prepare the public for situations in which additional information will be forthcoming or instructions could change over time(Vaughan & Tinker, 2009).

In addition to the names of the President and Vice President arising as the top words in our analysis, we observed other proper names commonly mentioned in news sources: Hanks (for actor Tom Hanks), Porter (for US Congresswoman Katie Porter) and Barack Obama (for the former US president). These names are illustrative for what they indicate the absence of – a designated spokesperson for public health, which is considered an essential part of a communication plan in emergencies and for which guidelines exist(Pan American Health Organization, 2009; US Department of Health and Human Services Centers for Disease Control and Prevention, 2018 Edition; World Health Organization, 2017). In pandemic situations, perception by the public that the designated spokesperson is trustworthy and credible is essential to their following instructions(World Health Organization, 2017). It is recommended that health experts work with communication specialists to improve response activities (Pan American Health Organization, 2009).

Given the numerous different news sources in the US each reaching some segment of the population, public health communication during a pandemic should be deliberate in the involvement of the media (US Department of Health and Human Services Centers for Disease Control and Prevention, 2018 Edition). A partnership with the news media is considered a best practice (Pan American Health Organization, 2009). Public health entities should also recognize the growing reliance of the public on social media for news requires communication plans be modernized. While websites can serve as a main repository for important information and updates during a pandemic, public health entities should also establish and maintain a presence on major social media sites such as Facebook, Twitter, Instagram, YouTube and Reddit, particularly since social media is increasingly where information is obtained by the public and circulated. An advantage to this approach is that public health entities have more control over content posted on their social media sites compared with news media sourcing information for their own coverage. However, a mere presence on these platforms is not enough; it is important to package public health information in appealing ways. Partnerships with social media personalities adept at creating viral information could achieve this.

Communication of important health information during times of communicable disease pandemics is crucial to informing and educating the public (Naik et al., 2019; US Department of Health and Human Services Centers for Disease Control and Prevention, 2018 Edition).

Particularly critical for public health efforts to control an outbreak and prevent additional cases is communication during initial stages of an epidemic as disease enters a population and begins to spread. Inherent challenges occur with communicating information during these initial stages as much about the agent (i.e., incubation period, routes of transmission) and disease (treatment approaches, severity) may be unknown, as was the case with COVID-19 caused by the novel coronavirus. As more is learned and knowledge grows while a pandemic unfolds, it is anticipated that health entities will provide additional information to the public and correct previously disseminated information as necessary(Vaughan & Tinker, 2009; World Health Organization, 2017). Modes of communication including a reliance on news and social media during a fast-moving pandemic should be nimble, flexible and efficient for this to be achieved (Federal Emergency Management Agency, December 2005).

## Data Availability

The data that support the findings of this study are available from the corresponding author (NG) upon reasonable request.

## Declaration Of Interest

### Authorship

Authors contributed to this work as follows: formulating the research question(s): WC, EA, TG, NMG; designing the study: WC, EA, TG; carrying it out: WC, EA; analyzing the data: WC, EA; writing the article: WC, EA, TG, NMG

